# Population pharmacokinetics of child-friendly dispersible amoxicillin/clavulanate tablets in African children with severe community acquired pneumonia

**DOI:** 10.64898/2026.09.18.26362138

**Authors:** Jennie van Dyk, Manna Semere Gebreyesus, Michelle Clements, Joseph F. Standing, David P. Moore, Ziyaad Dangor, Zainab Waggie, Ben Spittle, Veronica Mulenga, Chishala Chabala, Hilda Mujuru, W. Chris Buck, Muhammad Sidat, Jahit Sacarlal, Julia A. Bielicki, Victor Musiime, Hua Xu, Shabir A. Madhi, Mike Sharland, Paolo Denti, Roeland E. Wasmann, the PediCAP trial team

**Affiliations:** Division of Clinical Pharmacology, Department of Medicine, University of Cape Town, Cape Town, South Africa; Medical Research Centre Clinical Trials Unit at University College London, UK; School of Pharmacy, University College London, London, UK; Department of Pharmacy, Great Ormond Street Hospital for Children, London, UK; Department of Clinical Pharmacology, Stellenbosch University, South Africa; South Africa Medical Research Council Vaccines and Infectious Diseases Analytics Research Unit, Faculty of Health Science, University of the Witwatersrand, Johannesburg, South Africa; Department of Paediatrics and Child Health, Chris Hani Baragwanath Academic Hospital, and School of Clinical Medicine, Faculty of Health Sciences, University of the Witwatersrand, Johannesburg, South Africa; University Teaching Hospitals-Children’s Hospital, Lusaka, Zambia; School of Medicine, University of Zambia, Lusaka, Zambia; Department of Paediatrics and Child Health, University of Zimbabwe, Harare, Zimbabwe; Universidade Eduardo Mondlane, Faculdade de Medicina, Maputo, Mozambique; University of California Los Angeles, David Geffen School of Medicine, California Los Angeles, USA; Wits Infectious Diseases and Oncology Research Institute, Faculty of Health Sciences, University of the Witwatersrand, Johannesburg, South Africa; Centre for Neonatal and Paediatric Infection, City St George’s, University of London, UK; Department of Paediatrics and Child health, Makerere University Kampala, Uganda; Research Department, Joint Clinical Research Centre, Kampala, Uganda; Analytical Services International, City St George’s University of London, Cranmer Terrace, London

**Keywords:** Beta-lactam antibiotics, clavulanic acid, step-down therapy, pediatric dosing

## Abstract

**Background:** Severe community-acquired pneumonia (CAP) remains a leading cause of mortality in children under five. The PediCAP trial evaluated oral step-down therapy using dispersible amoxicillin or amoxicillin/clavulanate tablets in African children hospitalised with severe CAP. Dispersible tablet formulations are preferred in children; however, pharmacokinetic data for oral amoxicillin/clavulanate in paediatric populations remain limited. We characterised the pharmacokinetics of 4:1, 7:1, and 14:1 dispersible amoxicillin/clavulanate tablets in children enrolled in the PediCAP trial and predicted target attainment under different dosing schemes.

**Methods:** Children were randomised within 24 hours of starting intravenous antibiotics to oral step-down therapy with weight-banded dosing of twice-daily amoxicillin/clavulanate (4:1, 7:1, or 14:1). Plasma concentrations were quantified following intensive sampling and analysed using nonlinear mixed-effects modelling with allometric scaling and postmenstrual age-dependent clearance maturation. Pharmacodynamic indices included %fT>MIC for amoxicillin and %fT>C_t_ for clavulanate.

**Results:** A total of 157 children (median [range] age, 12 [2–70] months; weight, 8.4 [4.0-22.0] kg) contributed 707 paired concentrations. Amoxicillin was described by a two-compartment model and clavulanate by a one-compartment model, both with sequential zero-and first-order absorption. Typical clearance for an 8-kg individual at full maturation was 11.8L/h for amoxicillin and 6.1L/h for clavulanate. Pharmacokinetics were similar between the formulation ratios. Weight-banded dosing achieved balanced exposures across weight groups. Amoxicillin achieved 40%fT>MIC for MICs≤2mg/L. A proposed clavulanate target of 20%fT>0.5mg/L was achieved in most children even at the lowest amoxicillin/clavulanate ratio.

**Conclusion:** Weight-banded dosing using dispersible amoxicillin/clavulanate tablets provide consistent exposure profiles in African children with severe CAP.

**SUMMARY:** Population pharmacokinetic modelling of dispersible amoxicillin/clavulanate tablets (4:1, 7:1, 14:1) in 157 African children with severe pneumonia showed weight-banded dosing achieved consistent and adequate amoxicillin and clavulanate exposures across formulations.

## Introduction

Severe community acquired pneumonia (CAP) is the leading cause of childhood morbidity and hospitalisation worldwide, with the greatest burden in socioeconomically disadvantaged populations. It is also the deadliest infectious disease in children under five, claiming over 700,000 lives each year. Children in low-and lower-middle income countries (LMICs) account for 82% of global pneumonia deaths [1,2]. The World Health Organisation (WHO) recommends at least five days of intravenous antibiotics or continuation until clinical stability is achieved as treatment for severe paediatric CAP [3]. Consequently, management is associated with prolonged hospitalisation, increased healthcare and societal costs, acquisition of multidrug-resistant organisms, and heightened risk of nosocomial infection [1,4]. Rapid step-down to oral antibiotics, once clinical improvement is established, would enable earlier discharge from hospital and reduce these risks.

Amoxicillin is a first-line therapy for paediatric CAP and the most widely prescribed antibiotic in children [3,5]. Clavulanate, a β-lactamase inhibitor, can be administered alongside amoxicillin to protect against β-lactamase inhibition and extend its coverage to β-lactamase-producing organisms [6]. The recent PediCAP trial investigated oral step-down to amoxicillin or amoxicillin/clavulanate, using dispersible tablets, in African children hospitalised with severe CAP [7]. Amoxicillin/clavulanate is more typically prescribed in children as an oral suspension, but due to the instability of clavulanate in solution, this formulation requires cold-chain storage once reconstituted [8]. These requirements may significantly limit its use in resource-limited settings where refrigeration is not always available, risking drug degradation and compromised exposure [8,9]. Amoxicillin/clavulanate dispersible tablet formulations offer improved stability and are recognised as preferred formulations in children [10], but pharmacokinetic data for these formulations in children remain limited.

Amoxicillin and clavulanate are primarily eliminated by renal excretion with similar terminal half-lives of approximately 2 hours [11,12]. Amoxicillin demonstrates saturable absorption, with non-proportional increases in systemic exposure at daily doses above 1500 mg in adults [13,14]. Antibacterial activity is best characterised by the fraction of the dosing interval during which unbound concentrations exceed a target concentration (%fT>Ct), with approximately 20% associated with bacterial stasis and 40% with maximal bacterial killing [15,16].

While the pharmacokinetic effect of modifying amoxicillin dose levels has been well documented [14,17,18], the impact of varying clavulanate dose or ratios on systemic exposure remains poorly characterized. Both 4:1 and 7:1 amoxicillin/clavulanate dispersible tablets are included in the WHO Essential Medicines List for Children [5], but selection of the optimal amoxicillin/clavulanate ratio is limited by lack of clinical paediatric pharmacokinetic data for oral amoxicillin/clavulanate, especially for dispersible tablet formulations.

In this study, we aimed to characterise the pharmacokinetics of oral amoxicillin/clavulanate dispersible tablets in African children hospitalised with severe CAP and to explore the impact of varying amoxicillin/clavulanate ratios (4:1, 7:1, and 14:1) on systemic exposure.

## Methodology

### Study design and participants

The PediCAP trial [ISRCTN63115131] was a multicentre, open-label, parallel-group, 2 × 5 factorial randomised controlled trial with a parallel Phase II pharmacokinetic trial. It evaluated the impact of oral step-down to dispersible amoxicillin or amoxicillin/clavulanate tablets on effectiveness, safety, and selection of antibiotic resistance in severe childhood CAP. The study was conducted between April 2019 and March 2025 across nine centres in Mozambique, South Africa, Uganda, Zambia, and Zimbabwe.

Children aged 2 months to 6 years (weighing ≥3 to <30 kg) hospitalised with severe pneumonia and requiring at least 24 hours of intravenous antibiotic therapy were eligible. Recruitment was mandated to occur within 24 hours of commencement of intravenous antibiotic therapy. Exclusion criteria included known hypersensitivity to penicillins, requirement for non-protocol antibiotics, or receipt of long-term antibiotic therapy. Written informed consent was obtained from the parent or legal guardian, with additional consent obtained for participation in the pharmacokinetic sub-study. The study was approved by relevant ethics committees in the United Kingdom (ref: 16423 001), South Africa (ref: 190913B), Uganda (ref: 2019-62), Zambia (ref: 328-2019), Zimbabwe (ref: 221/19), and Mozambique (ref: 323/CNBS/22) and was conducted in accordance with the Declaration of Helsinki.

Full details of the PediCAP trial are available in the main paper [7]. Briefly, children who had received at least 24 hours of intravenous antibiotics were randomised to either step down to oral therapy once clinically stable and able to tolerate oral medication, or continue intravenous therapy. For the oral dosing, participants were randomised to either twice-daily oral amoxicillin or 7:1 amoxicillin/clavulanate. Those receiving the 7:1 amoxicillin/clavulanate were invited to participate in the pharmacokinetic sub-study. Additionally, in a parallel Phase II pharmacokinetic sub-study, children meeting the same eligibility criteria were randomised to oral step-down using either 4:1 or 14:1 amoxicillin/clavulanate for a total antibiotic duration of 6 days. Participants in the pharmacokinetic sub-studies were invited to undergo intensive pharmacokinetic sampling with the aim of enrolling 60 participants per group.

### Formulations and dosing

Oral step-down amoxicillin/clavulanate was administered twice-daily with dispersible tablets provided by Sandoz in three fixed-dose ratios: 4:1 (250/62.5 mg), 7:1 (200/28.5 mg), and 14:1 (150/10.725 mg) amoxicillin/clavulanate. Dose regimens were defined before the study and weight-banded according to **Table S1**. The selected doses accounted for both allometry and clearance maturation with age [19,20] to achieve similar exposure across weight bands. The dosing table was designed to keep the amoxicillin dose level as consistent as possible across cohorts, within the constraints of fixed tablet strengths, while modulating clavulanate amounts.

### Pharmacokinetic sampling and assay

Pharmacokinetic sampling was performed on the first observed morning dose of oral amoxicillin/clavulanate. Blood samples (ideally, but not invariably 2 mL each) were collected via an indwelling cannula immediately prior to, and 1, 2, 3, and 6 hours after amoxicillin/clavulanate administration in children weighing ≥5 kg. To comply with blood volume constraints in smaller children, for those 4-<5 kg and those <4kg, one and two samples, respectively, were randomly omitted amongst the 1, 2, 3 hours samples. Sampling times were determined using stochastic simulation and estimation (SSE), aiming to maximize information content while minimizing the proportion of post-dose samples falling below the limit of detection.

Samples were stored in EDTA tubes on ice immediately after blood draw and during transport to the laboratory for initial processing. EDTA blood was centrifuged at 1900 x g for 5 minutes and plasma stored as 2 x 0.5 mL aliquots in −80°C until shipment. All pharmacokinetic analyses were done at Analytical Services International Ltd (ASI), St. George’s Hospital, London, United Kingdom where plasma samples were quantified for amoxicillin and clavulanate using a validated Liquid Chromatography-Tandem Mass Spectrometry (LC-MS/MS) method. The limit of detection (LOD) was 0.05 and 0.005 mg/L for amoxicillin and clavulanate, respectively.

### Population pharmacokinetic modelling

Amoxicillin and clavulanate pharmacokinetic data were described simultaneously in plasma using nonlinear mixed effects modelling in NONMEM v7.5.1 (ICON Development Solutions, Ellicott City, MD, USA). Allometric scaling was applied *a priori* to clearance and volume, tested using either total body weight or fat-free mass as descriptors. Covariates were then investigated using a stepwise approach to describe variability in amoxicillin and clavulanate paediatric pharmacokinetic data. Full details of the population pharmacokinetic model development are provided in the **Supplementary Information (Section S2)**.

### Model-derived pharmacokinetics/pharmacodynamics

Individual pharmacokinetic estimates were obtained from the post-hoc step of the final model and used to derive amoxicillin and clavulanate exposures. Given the twice-daily dosing regimen, in which some weight bands received unequal morning and evening doses (e.g. 10-<14 kg receiving the 7:1 ratio, see **Table S1**), area under the curve is reported over a 24-hour dosing interval at steady-state (AUC_0-24, ss_) [Eq. 1] and compared with the mean AUC_0-24_ in adults receiving twice-daily 875/125 mg oral amoxicillin/clavulanate [14,17].

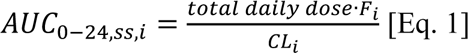

where total daily dose represents the total daily dose in mg of amoxicillin or clavulanate according to the weight-banded dosing regimen in **Table S1**, F_i_ is the individual relative bioavailability, and CL_i_ is the individual clearance. F_i_ was assumed the same across both dosing intervals and corresponds to the observed occasion.

Target attainment was evaluated for each participant over the dosing interval of the pharmacokinetic sampling day. For each participant, the percentage of the dosing interval during which free drug concentrations exceeded the minimum inhibitory concentration (%fT>MIC) was calculated for amoxicillin, and the percentage of the dosing interval during which free drug concentrations exceeded threshold concentrations (%fT>C_t_) was calculated for clavulanate. Free concentrations were derived assuming protein binding of 17% for amoxicillin and 22.3% for clavulanate [21,22].

For amoxicillin, %fT>MIC was evaluated across MIC values of 0.5, 1, 2, 4, and 8 mg/L, reflecting clinically relevant susceptibility breakpoints for a range of pathogens, as defined by CLSI and EUCAST [23,24]. For clavulanate, in the absence of a defined pharmacodynamic target, %fT>C_t_ was evaluated across a range of *in vitro* and *in vivo* threshold concentrations for beta-lactamase activity previously reported [16,23–28].

## Results

A total of 157 patients were included, with a median age of 13 (range 2−70) months and body weight of 8.4 (range 4.0−22.0) kg. Participant characteristics overall and by randomised amoxicillin/clavulanate ratio (4:1, 7:1, and 14:1) are summarised in **Table 1**. A total of 707 paired amoxicillin/clavulanate concentrations were available for analysis. Of these, 58 (8.20%) amoxicillin and 114 (16.1%) clavulanate concentrations were below the LOD, and all but one were observed at the pre-dose sample. Twenty-four (15%) participants received their first oral dose of the step-down phase at the time of sampling.

**Table 1.**
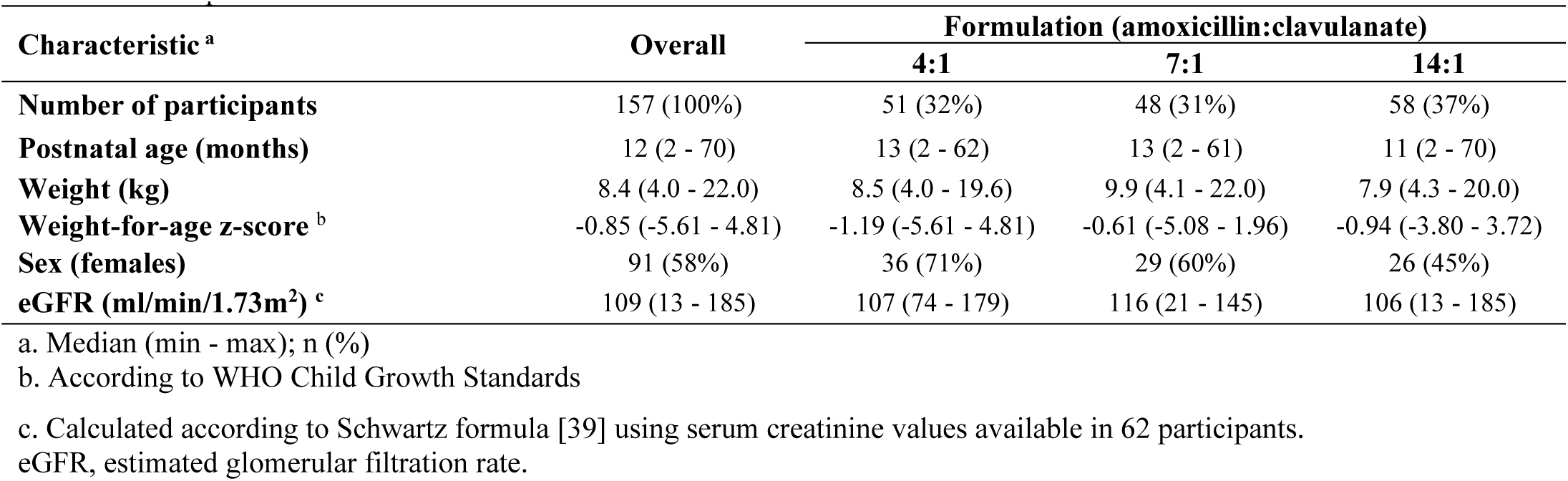
Participant characteristics.

### Population pharmacokinetic modelling

Data were best described by a two-compartment model with sequential zero-and first-order absorption for amoxicillin, and a one-compartment model with lag time and sequential zero-and first-order absorption for clavulanate. To account for high variability in pre-dose concentrations, a scaling factor was applied as a fixed effect to inflate the between-occasion variability on bioavailability (dOFV = −24.8; P < 0.001) for concentrations measured after an unobserved dose. Correlations between amoxicillin and clavulanate random effects were identified for bioavailability (r = 0.658 [dOFV = −88.96; P<0.001]) and zero-and first-order absorption parameters (r = 0.740 [dOFV = −70.91; P<0.001] and 0.866 [dOFV = −41.37; P<0.001], respectively), corresponding to moderate to strong correlations, and were retained in the combined model.

Allometric scaling using total body weight improved the model fit (dOFV = −43.81) and was similar to using fat-free mass (dOFV = −1.62). Total body weight was therefore retained to describe amoxicillin and clavulanate disposition. Including the effect of maturation with age on clearance significantly improved model fit (dOFV = −41.9; P < 0.001, compared to model without clearance maturation). The maturation function, displayed in **Figure S1**, had a Hill coefficient of 3.42 (95%CI: 3.06-3.80), with clearance expected to reach 50% maturity at a postmenstrual age of 44.9 (95%CI: 42.2-47.2) weeks.

In participants where serum creatinine data was available, we observed a directly proportional trend between estimated glomerular filtration rate (eGFR) and amoxicillin and clavulanate clearance. We attempted to test this effect in the model; however, due to the large proportion of participants with missing serum creatinine data (61%), the difficulty of imputing missing serum creatinine values in this population, and the resulting model instability, including eGFR became impractical and was not retained the final model. The typical clearance for an 8 kg individual, assuming full maturation, was estimated at 11.8 (95%CI: 10.9−12.7) L/h for amoxicillin and 6.10 (95%CI: 5.52−6.82) L/h for clavulanate.

No other significant differences in pharmacokinetics were identified between the 4:1, 7:1, and 14:1 formulations, and no effect of weight-for-age z-scores or dose was observed. Final model parameter estimates with 95% confidence intervals are presented in **Table 2**. Visual predictive checks demonstrated that the final model adequately described amoxicillin and clavulanate concentrations across all formulation ratios, as seen in **Figure 1**.

**Figure 1.**
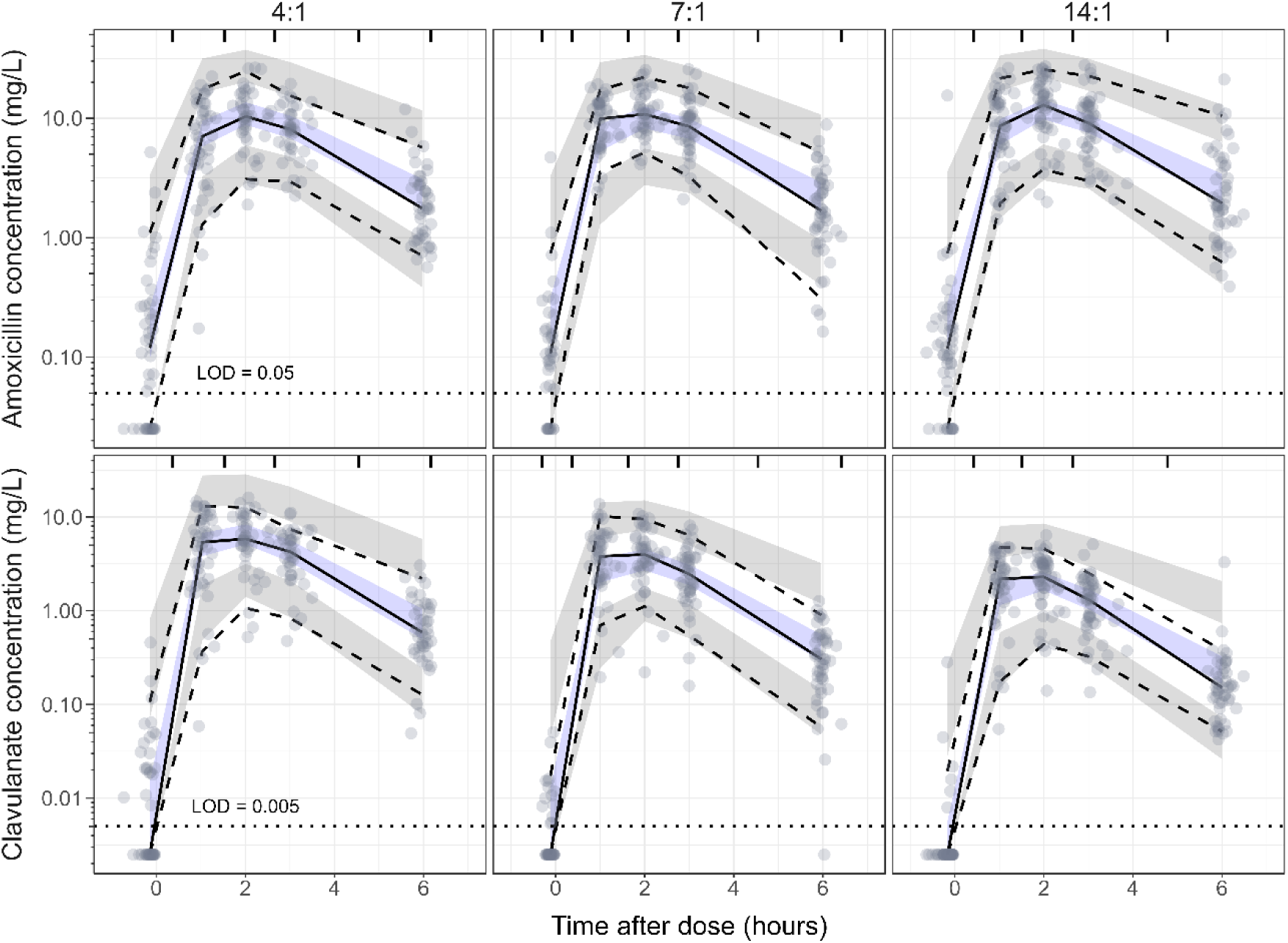
Visual predictive check for amoxicillin (top panel) and clavulanate (bottom panel) across amoxicillin/clavulanate formulation. Solid circles represent observed data. The dashed (lower), solid, and dashed (upper) lines represent the 5th, 50th, and 95th percentiles of observed data. The shaded areas represent the model-predicted 95% confidence intervals for the same percentiles. Top-axis tick marks indicate bin boundaries. Dotted line represents the assay limit of detection (LOD) for amoxicillin (0.05 mg/L) and clavulanate (0.005 mg/L). Values below the LOD were imputed to half the LOD for the visualization purposes.

**Table 2.**
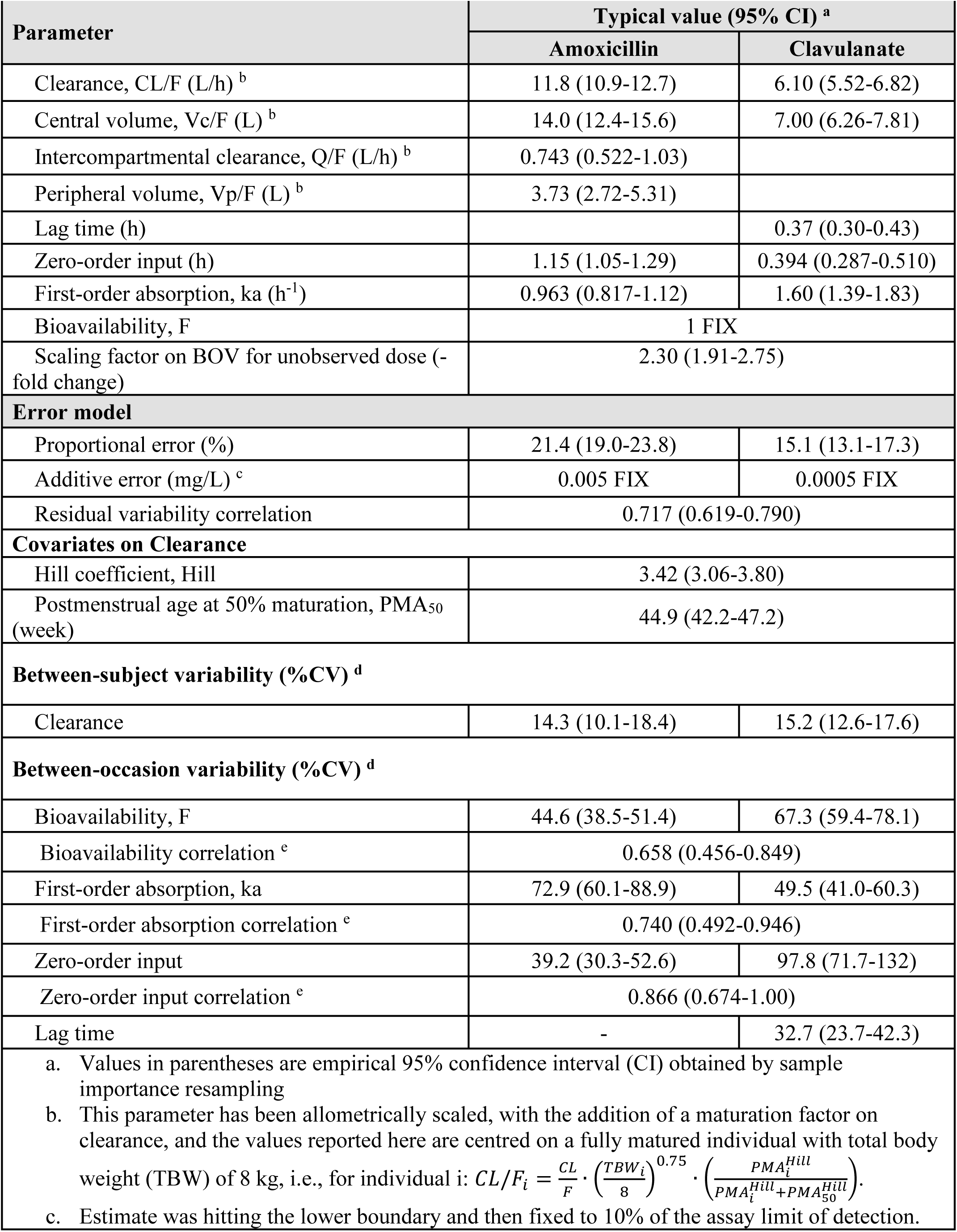
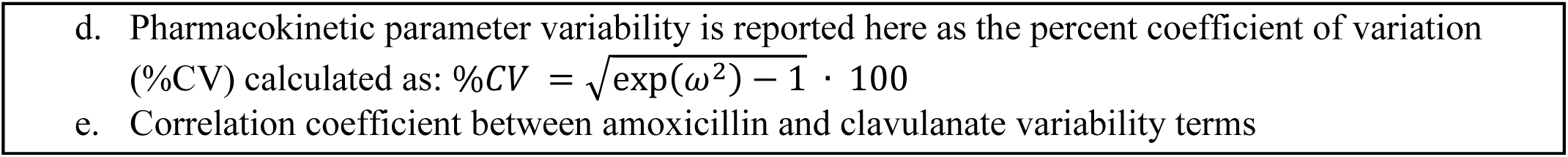
Population pharmacokinetic model parameter estimates.

| Parameter | Typical value (95% CI) <sup>a</sup> |  |
| --- | --- | --- |
|  | Amoxicillin | Clavulanate |
| Clearance, CL/F (L/h) <sup>b</sup> | 11.8 (10.9-12.7) | 6.10 (5.52-6.82) |
| Central volume, Vc/F (L) <sup>b</sup> | 14.0 (12.4-15.6) | 7.00 (6.26-7.81) |
| Intercompartmental clearance, Q/F (L/h) <sup>b</sup> | 0.743 (0.522-1.03) |  |
| Peripheral volume, Vp/F (L) <sup>b</sup> | 3.73 (2.72-5.31) |  |
| Lag time (h) |  | 0.37 (0.30-0.43) |
| Zero-order input (h) | 1.15 (1.05-1.29) | 0.394 (0.287-0.510) |
| First-order absorption, ka (h<sup>-1</sup>) | 0.963 (0.817-1.12) | 1.60 (1.39-1.83) |
| Bioavailability, F | 1 FIX |  |
| Scaling factor on BOV for unobserved dose (-fold change) | 2.30 (1.91-2.75) |  |
| Error model |  |  |
| Proportional error (%) | 21.4 (19.0-23.8) | 15.1 (13.1-17.3) |
| Additive error (mg/L) <sup>c</sup> | 0.005 FIX | 0.0005 FIX |
| Residual variability correlation | 0.717 (0.619-0.790) |  |
| Covariates on Clearance |  |  |
| Hill coefficient, Hill | 3.42 (3.06-3.80) |  |
| Postmenstrual age at 50% maturation, PMA<sub>50</sub> (week) | 44.9 (42.2-47.2) |  |
| Between-subject variability (%CV) <sup>d</sup> |  |  |
| Clearance | 14.3 (10.1-18.4) | 15.2 (12.6-17.6) |
| Between-occasion variability (%CV) <sup>d</sup> |  |  |
| Bioavailability, F | 44.6 (38.5-51.4) | 67.3 (59.4-78.1) |
| Bioavailability correlation <sup>e</sup> | 0.658 (0.456-0.849) |  |
| First-order absorption, ka | 72.9 (60.1-88.9) | 49.5 (41.0-60.3) |
| First-order absorption correlation <sup>e</sup> | 0.740 (0.492-0.946) |  |
| Zero-order input | 39.2 (30.3-52.6) | 97.8 (71.7-132) |
| Zero-order input correlation <sup>e</sup> | 0.866 (0.674-1.00) |  |
| Lag time | - | 32.7 (23.7-42.3) |
| a. Values in parentheses are empirical 95% confidence interval (CI) obtained by sample importance resampling<br>b. This parameter has been allometrically scaled, with the addition of a maturation factor on clearance, and the values reported here are centred on a fully matured individual with total body weight (TBW) of 8 kg, i.e., for individual i: $CL/F_i = \frac{CL}{F} \cdot \left(\frac{TBW_i}{8}\right)^{0.75} \cdot \left(\frac{PMA_i^{Hill}}{PMA_i^{Hill} + PMA_{50}^{Hill}}\right)$.<br>c. Estimate was hitting the lower boundary and then fixed to 10% of the assay limit of detection. |  |  |

- d. Pharmacokinetic parameter variability is reported here as the percent coefficient of variation (%CV) calculated as: $\%CV = \sqrt{\exp(\omega^2) - 1} \cdot 100$ - e. Correlation coefficient between amoxicillin and clavulanate variability terms

### Exposure metrics

Model-derived pharmacokinetic and pharmacodynamic parameters for twice-daily amoxicillin/clavulanate in participants are summarised in **Table 3** across 4:1, 7:1, and 14:1 formulations. Geometric mean amoxicillin AUC_0–24,*ss*_ was similar across formulations, and clavulanate AUC_0–24,*ss*_ decreased with increasing amoxicillin/clavulanate ratio (i.e. decreasing clavulanate content). One participant receiving the 14:1 formulation exhibited markedly elevated exposures (amoxicillin AUC_0–24,*ss*_: 135 mg·h/L vs geometric mean 77.8 mg·h/L; clavulanate AUC_0–24,*ss*_: 31.3 mg·h/L vs geometric mean 10.3 mg·h/L). Among participants with available serum creatinine data, this individual had reduced renal function, with an estimated eGFR of 13 mL/min/1.73m^2^. Overall, exposures appeared balanced across weight groups and were above the adult reference, as seen in **Figure 2**.

**Figure 2.**
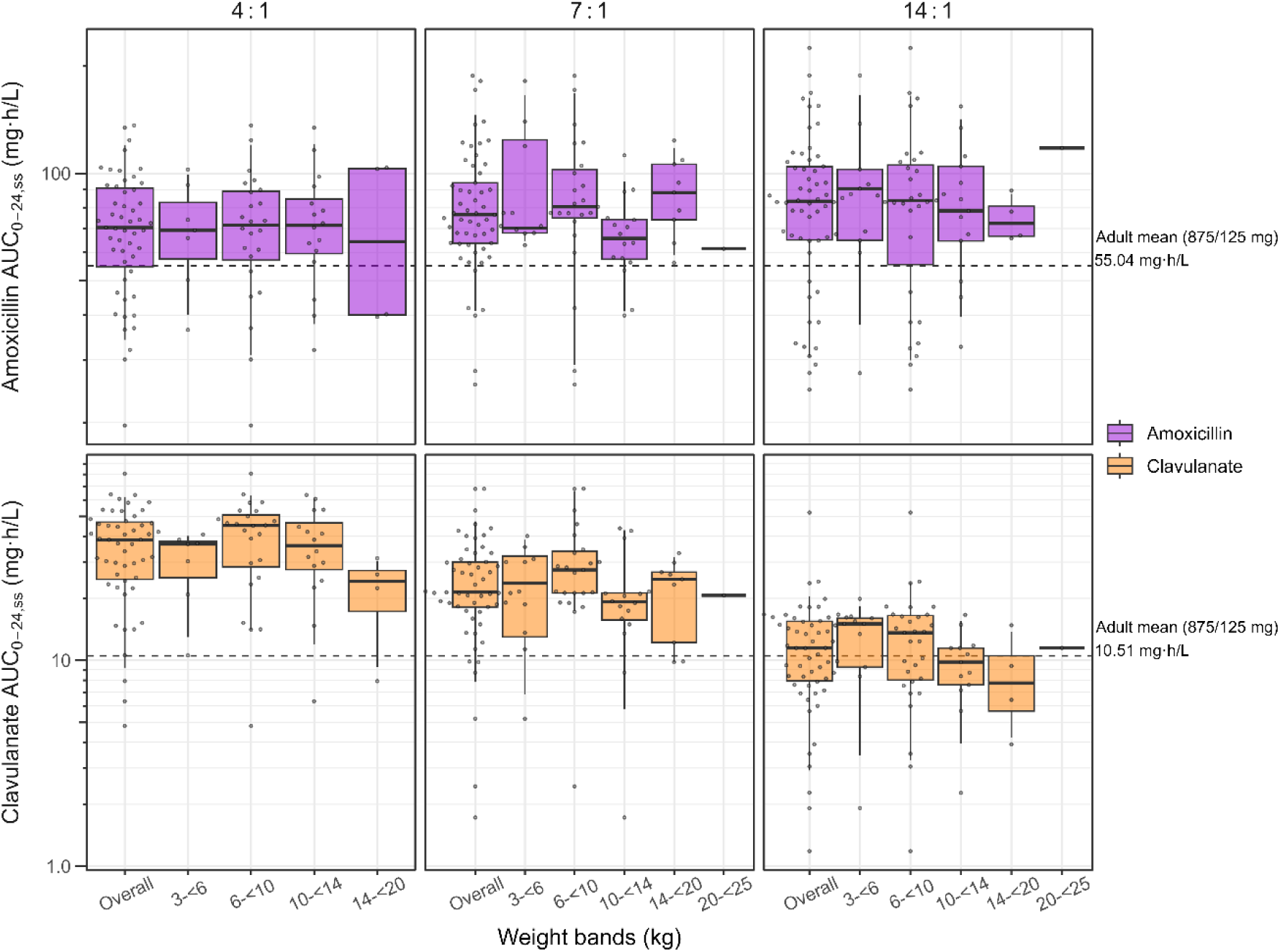
Amoxicillin and clavulanate exposure using weight banded dosing. Model-derived AUC_0-24_ of amoxicillin (top panel) and clavulanate (bottom panel) for each participant (solid circles) across weight bands receiving 4:1, 7:1, and 14:1 amoxicillin/clavulanate dispersible tablets. The whiskers represent the 5^th^ and 95^th^ percentiles. Dashed lines indicate adult reference mean AUC_0-24_ values for amoxicillin [17] and clavulanate [14], respectively, after twice-daily oral 875/125 mg amoxicillin/clavulanate in adults.

**Table 3.** Model-derived pharmacokinetic (AUC_0-24, ss_, C_avg_, t_1/2_) and pharmacodynamic (%fT > MIC) values of twice-daily oral amoxicillin/clavulanate in study participants.

| Parameters <sup>a</sup> | Formulation (amoxicillin:clavulanate) |  |  |
| --- | --- | --- | --- |
|  | 4:1 | 7:1 | 14:1 |
| **Amoxicillin** |  |  |  |
| AUC<sub>0-24, ss</sub> (mg·h/L) | 67.2 (42.5) | 77.7 (41) | 77.8 (53.8) |
| C<sub>avg</sub> (mg/L) | 2.8 (42.5) | 3.24 (41) | 3.24 (53.8) |
| t<sub>1/2</sub> (h) <sup>b</sup> | 3.93 (3.3 - 4.85) | 4.05 (3.34 - 4.95) | 3.88 (3.44 - 4.83) |
| %fT > MIC (mg/L) <sup>d</sup> |  |  |  |
| MIC = 0.5 | 66.2 (46.8 - 100) | 63.6 (34.7 - 97) | 66.7 (42.9 - 100) |
| MIC = 1 | 53.1 (34.3 - 96.8) | 51.2 (27.1 - 95.3) | 53.7 (33.2 - 97.8) |
| MIC = 2 | 40.2 (19.1 - 82.9) | 39.3 (18.1 - 73.8) | 42.9 (22.4 - 88.1) |
| MIC = 4 | 27.3 (0 - 59.6) | 27.4 (6.45 - 49.3) | 30.9 (0 - 61.9) |
| MIC = 8 | 10.1 (0 - 33.2) | 11.7 (0 - 31.5) | 17.1 (0 - 43) |
| **Clavulanate** |  |  |  |
| AUC<sub>0-24, ss</sub> (mg·h/L) | 31.8 (66.4) | 21.3 (74.5) | 10.3 (71.5) |
| C<sub>avg</sub> (mg/L) | 1.33 (66.4) | 0.888 (74.5) | 0.428 (71.5) |
| t<sub>1/2</sub> (h) <sup>c</sup> | 0.94 (0.672 - 1.31) | 0.914 (0.571 - 1.13) | 0.912(0.712 - 1.21) |

- a. Geometric mean (%CV), median (min - max) - b. Calculated using $t_{1/2} = \frac{\ln 2}{0.5 \left[ \left( \frac{CL}{V_c} + \frac{Q}{V_c} + \frac{Q}{V_p} \right) - \sqrt{\left( \frac{CL}{V_c} + \frac{Q}{V_c} + \frac{Q}{V_p} \right)^2 - 4 \cdot \left( \frac{CL}{V_c} \right) \cdot \left( \frac{Q}{V_p} \right)} \right]}$ - c. Calculated using $t_{1/2} = \frac{\ln 2}{CL/V_c}$ - d. Unbound concentration calculated assuming 17% serum protein binding [22]
AUC<sub>0-24, ss</sub>, area under the concentration-time curve over 24-hour dosing interval at steady-state; C<sub>avg</sub>, average drug concentration over 24-hour dosing interval; t<sub>1/2</sub>, terminal half-life; MIC, minimum inhibitory concentration; %fT>MIC, percentage of dosing interval where unbound concentrations exceed MIC.

As expected, amoxicillin target attainment over the dosing interval on the sampling day was comparable across the three tablet formulations. The commonly used efficacy target of 40%fT>MIC was achieved at MIC≤2 mg/L. At higher MIC values, %fT>MIC declined below 40%, with lower bounds approaching zero. Clavulanate pharmacodynamic target attainment is plotted in **Figure 3** and differs by formulation ratio. The suggested threshold target for clavulanate of 20%fT>C_t_ [16] was achieved most consistently with the 4:1 formulation, where median %fT>C_t_ exceeded 20% at C_t_≤2.0 mg/L. The 7:1 formulation achieved 20%fT>C_t_ at C_t_ ≤1.0 mg/L, whereas the 14:1 formulation met the threshold only at lower C_t_ values (≤0.5 mg/L).

**Figure 3.**
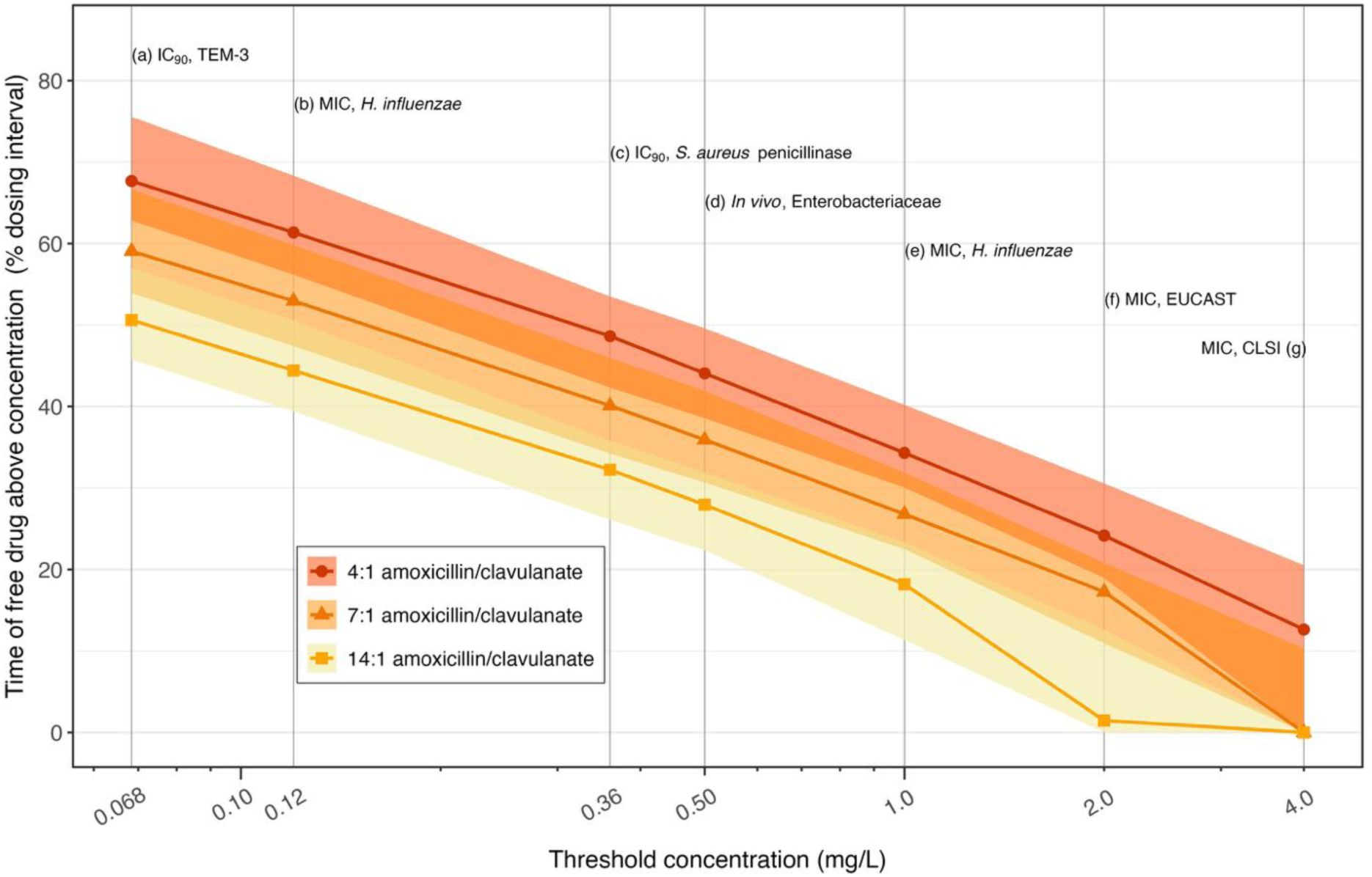
Clavulanate %fT>C_t_ by amoxicillin/clavulanate formulation ratio. The median (solid line) and interquartile range (shaded area) of the dosing interval where unbound clavulanate concentrations exceed predefined threshold concentrations (%fT>C_t_) were derived from individual post-hoc pharmacokinetic parameter estimates in study participants receiving 4:1 (circles), 7:1 (triangles), and 14:1 (squares) oral amoxicillin/clavulanate dispersible tablets. Grey vertical lines denote threshold concentrations derived from literature: (a) IC₉₀ for inhibition of TEM-3 β-lactamase [28]; (b) fixed concentration at which 74% and 91% of *Haemophilus influenzae* isolates were susceptible at 0.5 and 1 mg/L amoxicillin, respectively [27]; (c) IC₉₀ for inhibition of *Staphylococcus aureus* penicillinase [26]; (d) plasma concentration associated with β-lactamase inhibition in Enterobacteriaceae, derived from murine infection model [16]; (e) revised susceptible breakpoint for *H. influenzae* (amoxicillin/clavulanate 2/1 mg/L) [25]; (f) EUCAST fixed clavulanate concentration for amoxicillin susceptibility testing [23]; and (g) CLSI resistance breakpoint for *S. aureus*, *H. influenzae*, and *Streptococcus pneumoniae* (amoxicillin–clavulanate 8/4 mg/L) [24].

## Discussion

In this study, we described the pharmacokinetics of child-friendly dispersible amoxicillin/clavulanate tablets (4:1, 7:1, and 14:1) in a large cohort of young African children with severe community-acquired pneumonia. Weight-banded dosing, designed to account for allometric scaling and maturation of clearance with age, achieved balanced amoxicillin and clavulanate exposures across weight groups. A single maturation function, closely resembling GFR maturation, adequately described changes in clearance with age for both compounds. No clinically meaningful differences in pharmacokinetics were observed between formulation ratios, and clavulanate demonstrated dose-proportional pharmacokinetics across the dose range studied. Doses achieved adequate exposures in most participants, even at the amoxicillin/clavulanate ratio with the lowest clavulanate dose.

Paediatric dosing of antibiotics has historically relied on linear scaling with body weight, i.e. the same mg/kg dose as in adults. This approach makes the implicit assumption that drug clearance increases and decreases linearly with body size. This is despite the well-established theory of allometry, which has shown that drug clearance, along with other metabolic processes, scales with body size with an exponent of 3/4 [29]. Another important factor influencing clearance besides body size is age, which has an important effect in the first few years of life, when elimination organs and pathways are immature and therefore not fully functional [29]. A fixed mg/kg dose ignores this nonlinearity in the relationship with body size and the process of maturation, thus systematically under-or over-estimating the total dose required in children at the extremes of the studied age and weight range. This risks sub-or supra-therapeutic (and potentially toxic) exposure in the youngest, smallest children in particular [19].

The weight-banded dosing regimen used in this study was designed to account for both processes simultaneously, combining allometric scaling with the expected maturation trajectory of clearance with age. The results show that it achieved balanced amoxicillin and clavulanate exposure across all weight groups, avoiding the systematic exposure gradient typically produced by mg/kg dosing [19] and ensuring children across weights and ages receive comparable antibiotic exposure. This finding provides empirical support for the growing shift toward allometry-and maturation-informed weight-banded dosing in children, which is increasingly adopted by regulators and guideline groups to simplify prescribing and reduce dosing error [30].

While the effect of body size is predictable and consistent across many drugs, the maturation of clearance can significantly vary due to different elimination mechanisms and typically reflects the developmental trajectory of the pathways involved [29]. In this study, amoxicillin and clavulanate followed a common maturation profile that closely mirrored GFR development [31], reaching approximately 90% of mature function by one year of age in term-born infants. Because clearance continues to change substantially up to this age, maturation remains an important determinant of disposition during infancy, after which body size becomes the predominant driver of clearance and allometric scaling alone can reasonably guide dosing. A shared maturation profile between amoxicillin and clavulanate also benefits their co-formulation, as it ensures that the effective amoxicillin/clavulanate ratio delivered to the patient remains stable as children grow and develop.

Amoxicillin has previously been reported to show less-than-proportional increases in exposure at higher doses, an effect attributed to saturable absorption via carrier-mediated intestinal uptake [17]. Because the amoxicillin dose level was held approximately constant across the three amoxicillin/clavulanate ratios in this study, no formulation effect on amoxicillin pharmacokinetic parameters was identified, and this saturability could not be evaluated. In contrast, increasing the clavulanate dose across formulations resulted in proportional increases in exposure, indicating linear pharmacokinetics over the dose range studied. This is consistent with reports that clavulanate, a smaller molecule than amoxicillin, is absorbed predominantly by passive diffusion rather than a saturable carrier-mediated process [32–34]. At the studied doses, modifying the clavulanate dose to achieve different amoxicillin-to-clavulanate ratios is unlikely to alter clavulanate pharmacokinetics beyond the expected, proportional increase in exposure.

Direct comparison with previous pharmacokinetic studies is complicated by differences in study populations, treatment, and model standardisation. Detailed standardised comparisons are provided in the **Supplementary Information (Section S3, Tables S2–S3).** The most notable finding was that our apparent clearance and volume estimates for amoxicillin were consistently higher (2-to 7-fold) than previously reported in oral studies [12,17,35,36], whereas the discrepancy for clavulanate was smaller (0.43-to 3-fold) [14,18,36]. The larger discrepancy for amoxicillin may reflect differences in apparent bioavailability at the higher doses used in this study, driven by the reported saturability of amoxicillin absorption [13,17]. extrapolation between studies conducted at substantially different amoxicillin dose levels should therefore be interpreted with caution. This pattern was less evident for clavulanate despite also being given at relatively higher doses than in comparator studies, which is consistent with the dose proportionality observed across the studied dose range.

Maturation of amoxicillin and clavulanate clearance has been described previously. Barker *et al.* [12] analysed amoxicillin pharmacokinetics in neonates up to children under 16 years old and reported 50% maturation of clearance at a postmenstrual age of 42.6 weeks (Hill coefficient 2.68), broadly consistent with our estimate of 44.9 weeks (Hill coefficient 3.42). Their analysis also incorporated a postnatal age covariate to capture the rapid changes in clearance occurring within the first days of life. Schouwenberg *et al.* [18] similarly identified postnatal age-related changes in clavulanate clearance in preterm and term neonates during the first year of life, which we were unable to characterise since our study did not include neonates.

Target attainment analyses showed that most participants achieved pharmacodynamic targets for amoxicillin up to an MIC of 2 mg/L, which exceeds previously reported thresholds for twice-daily oral dosing in adults [14,17]. For clavulanate, we report target attainment at several concentrations (Ct) and thresholds (%fT>Ct), since no clear target has been established. Target attainment of the 4:1 and 7:1 amoxicillin/clavulanate formulations were comparable to those previously reported in neonates and infants [18]. Some studies report that lower thresholds of 20%fT>0.5 mg/L are sufficient for Enterobacteriaceae β-lactamase inhibition [16], which were achieved in the majority of participants even at the ratio with the lowest clavulanate dose (14:1 ratio). However, interpretation remains limited by the lack of consensus on optimal clavulanate thresholds.

Oral amoxicillin/clavulanate remains a first-choice antibiotic in children for several indications, including lower urinary tract infections, acute sinusitis, low-risk febrile neutropenia, and skin and soft tissue infections, and a second-choice option for osteomyelitis, acute otitis media, and surgical prophylaxis [5]. Although this study evaluated treatment of severe community-acquired pneumonia, the pharmacokinetic findings are applicable to other indications. As demonstrated by our predictions, the optimal dose, amoxicillin/clavulanate ratio, and pharmacodynamic target for any given indication will depend on the likely pathogen and its susceptibility profile, for which this population pharmacokinetic model provides a tool to inform such dosing decisions.

This study has some limitations. Pharmacokinetic sampling was relatively sparse (3 to 5 samples per participant) due to the practical constraints of repeated blood sampling in young children. However, we used nonlinear mixed-effects modelling to interpret the data, which “borrows” information across the population and is known to be robust against limited (sparse) information in individual participants [37]. Serum creatinine information was only available in a small number of children, so we were unable to incorporate renal function as a covariate. Using age-based maturation adequately described the development of renal clearance in this population, but caution is warranted when applying these findings to children with significantly impaired renal function, in whom exposures may be substantially elevated. Additionally, the maturation function was developed using data from the age range represented in this study. its applicability to neonates, in whom rapid postnatal physiological changes may not be fully captured, is therefore uncertain [38]. Finally, interpretation of clavulanate target attainment remains limited by the absence of established pharmacodynamic targets linked to clinical outcomes.

In conclusion, this study provides important pharmacokinetic data for child-friendly dispersible amoxicillin/clavulanate formulations in young African children. Weight-banded dosing, adjusted for allometric scaling with body weight and maturation, achieved balanced amoxicillin and clavulanate exposures across weight groups, producing more consistent exposure profiles than expected from fixed mg/kg dosing. Clearance maturation closely followed renal development, with body size becoming the predominant determinant after the first year of life. Applied with pathogen-specific pharmacodynamic targets, the pharmacokinetic model developed here can inform dosing strategies for amoxicillin/clavulanate across the range of indications for which it is used in children.

## Data Availability

All data produced in the present study are available upon reasonable request to the authors

## Acknowledgements

We thank the Mozambican, South African, Ugandan, Zambian, and Zimbabwean trial sites and the children and caregivers for participating in the PediCAP trial.

We would like to acknowledge Michael Borek for expert discussions on amoxicillin/clavulanate pharmacokinetics. Sandoz provided oral amoxicillin and amoxicillin/clavulanate fixed dose combination dispersible tablets but had no role in the design or conduct of the trial. The amoxicillin 200 mg/clavulanate 28.5 mg 7:1 novel dispersible tablet used in the trial was included in the 2023 WHO Essential Medicines List for Children following a submission from Sandoz. MS Chaired the 2023 WHO Expert Committee on Selection and Use of Essential Medicines.

We would also like to acknowledge the ICTS High Performance Computing team at the University of Cape Town (https://ucthpc.uct.ac.za/) for providing us with the resources to perform the calculations in this study.

This project is part of the EDCTP2 programme supported by the European Union (grant number RIA2017MC-2023).

## Author contributions

M. Se. G., M. C., J. F. S., D. P. M., B. S., V. Mulenga, C. C., H. M., W. C. B., J. S., J. A. B., V. Musiime, H. X., S. A. M., M. Sh., P. D., and R. E. W. contributed to the study conception and design. M. Se. G., M. C., D. P. M., B. S., V. Mulenga, C. C., H. M., W. C. B., J. S., J. A. B., V. Musiime, H. X., S. A. M., and M. Sh. contributed to the acquisition of data. J. van D., M. Se. G, P. D., and R. E. W. contributed to the analysis of data. J. van D. wrote the initial draft of the manuscript. All authors helped interpret the findings and reviewed and approved the final version of the manuscript. The complete listing of the PediCAP trial study team is included within **Supplementary Information (Section S1).**

## Notes

### Clinical Trial

ISRCTN63115131

### Clinical Protocols

https://www.thelancet.com/cms/10.1016/S0140-6736(26)00879-2/attachment/a2917fa1-b16b-4bd6-8e81-1a73818d2f0a/mmc1.pdf

### Author Declarations

Local ethical approval was sought and obtained in each country, specifically Mozambique, Comite Nacional de Bioethica Para a Saude (IRB00002657) REF 323/CNBS/22; South Africa, University of the Witwatersrand Human Research Ethics Committee (Medical) REF 190913B; Uganda, Makarere University Research Ethics Committee REF 2019-162; Zambia, University of Zambia Biomedical Research Ethics Committee REF 328-2019; Zimbabwe, Joint Research Ethics Committee for the University of Zimbabwe, College of Health Sciences and Parirenyatwa Group of Hospitals REF 221/19; UK, University College London Research Ethics Committee REF 16423/001.

