## supplementary information for "Population pharmacokinetics of child-friendly dispersible amoxicillin/clavulanate tablets in African children with severe community acquired pneumonia"

**TARGET JOURNAL**: CID

**AUTHOR LIST**

Jennie van Dyk* (1), Manna Semere Gebreyesus (1), Michelle Clements (2), Joseph F. Standing (3, 4, 5), David P. Moore (6, 7), Ziyaad Dangor (6), Zainab Waggie (7), Ben Spittle (2), Veronica Mulenga (8, 9), Chishala Chabala (8, 9), Hilda Mujuru (10), W. Chris Buck (11, 12), Muhammad Sidat (11), Jahit Sacarlal (11), Julia A. Bielicki (14), Victor Musiime (15, 16), Hua Xu (17), Shabir A. Madhi (6, 13), Mike Sharland (14), Paolo Denti (1), Roeland E. Wasmann (1), the PediCAP trial team**.

(1) Division of Clinical Pharmacology, Department of Medicine, University of Cape Town, Cape Town, South Africa

(2) Medical Research Centre Clinical Trials Unit at University College London, UK

(3) School of Pharmacy, University College London, London, UK

(4) Department of Pharmacy, Great Ormond Street Hospital for Children, London, UK

(5) Department of Clinical Pharmacology, Stellenbosch University, South Africa

(6) South Africa Medical Research Council Vaccines and Infectious Diseases Analytics Research Unit, Faculty of Health Science, University of the Witwatersrand, Johannesburg, South Africa

(7) Department of Paediatrics and Child Health, Chris Hani Baragwanath Academic Hospital, and School of Clinical Medicine, Faculty of Health Sciences, University of the Witwatersrand, Johannesburg, South Africa

(8) University Teaching Hospitals-Children's Hospital, Lusaka, Zambia

(9) School of Medicine, University of Zambia, Lusaka, Zambia

(10) Department of Paediatrics and Child Health, University of Zimbabwe, Harare, Zimbabwe

(11) Universidade Eduardo Mondlane, Faculdade de Medicina, Maputo, Mozambique

(12) University of California Los Angeles, David Geffen School of Medicine, California Los Angeles, USA

(13) Wits Infectious Diseases and Oncology Research Institute, Faculty of Health Sciences, University of the Witwatersrand, Johannesburg, South Africa

(14) Centre for Neonatal and Paediatric Infection, City St George's, University of London, UK

(15) Department of Paediatrics and Child health, Makerere University Kampala, Uganda

(16) Research Department, Joint Clinical Research Centre, Kampala, Uganda

(17) Analytical Services International, City St George's University of London, Cranmer Terrace, London

**Members listed in the **supplementary information (Section S1).**

### Supplementary text

#### Section S1: List of investigators

##### Participating Sites

Mozambique:

Universidade Eduardo Mondlane, Faculdade de Medicina, Maputo: Cláudia Massitela, Winete Joaquim, Alice Maieca, Uneisse Cassia, Quim Adriano, Elias Manjate, Sonia Martins, Alfeu Passanduca.

Hospital Geral de Mavalane, Maputo: Nádia Manjate, Mara Bombe.

Hospital Provincial da Matola, Matola: Eleutéria Macanze, Sheila Nhacumba.

Hospital Geral José Macamo, Maputo: Faiaz Issa, Natércia Florindo.

Hospital Central de Maputo, Maputo: Josina Chilundo, Ana Sigaúque, Rosalina Chilengue.

South Africa:

Africa Health Research Institute, Durban: Nigel Klein, Leatitia Kampiire, Anne Derache, Samkelisiwe Buthelezi, Senamile Makari.

The Chris Hani Baragwanath Hospital, Johannesburg: Sandisiwe Nkosi, Sihle Zindela, Nomhle Nkumana, Ntombi Khumalo, Lethabo Nkosi, Tshepiso Msibi, Brandon Jones, Lora Frank.

Zambia:

University Teaching Hospital Zambia, Lusaka: Monica Kapasa, Gae Mundundu, Patricia Ngoma, Jacqueline H. Kaira, Oliver Mwenechanya, Anne Chimolula, Bwalya Simunyola, Martha Sakala, Joyce C. Lungu, Chimuka Kwale, Dorothy Zangata, Naomi Mumba.

Chawama General Hospital Zambia, Lusaka: Jacqueline Mulako, Mundimba Makwamba, Frederick Chansa, Carolyn Zulu.

Zimbabwe:

University of Zimbabwe Faculty of Medicine and Health Sciences, Clinical Research Centre, Harare: Kusum J. Nathoo, Ennie Chidziva, Shepherd Mudzingwa, Misheck Phiri, Ruth Nhema, Godfrey Musoro, Vivian Mumbiro, Moses Chitsamatanga, Joy Chimhanzi, Shirley Mutsai, Dorcas Murungu, Wendy Mapfumo.

**Trial coordination and conduct**

Fondazione Penta, Padova, Italy: Musakanya Ching'andu, Francesca Viero.

University of Cape Town, Cape Town, South Africa: Susan Cleary, Lucy Cunnama, Mutsawashe Chitando, Paolo Denti, Roeland Wasmann, Manna Semere Gebreyesus, Jennie van Dyk.

Swiss Tropical and Public Health Institute, Basel, Switzerland: Günther Fink, Gillian Levine.

Global Antibiotic Research and Development Partnership, Geneva, Switzerland: Sally Ellis.

Centre for Neonatal and Paediatric Infection, St George's, University of London, UK: Kamla Pillay, Louise Hill, Tatiana Munera-Huertas, Jan Goelen.

Medical Research Council Clinical Trials Unit at University College London, London, UK: Clare Shakeshaft, Farjana Haque, Hanh Nguyen, Emily Dennis, Tatiana Sarfati, Brendan Murphy, Kolpona Begum, Nishdha Naufal, Hannah Sweeney, Jemima Shickle, Fatima Mohamed, Lee Barker, Marion Hall, Mallory Williams, Charlotte Hartley, Annabelle South.

**Trial oversight**

Trial Steering Committee members: Elizabeth Molyneux (Chair), Elizabeth Obimbo, Shamim Qazi, Somwe Wa Somwe, Robin Green.

Data Monitoring Committee members: Mainga Hamaluba, Tim Peto (Chair), Andrew Prendergast, Haroon Saloojee, Margaret Siwale.

Endpoint Review Committee members: Felicity Fitzgerald, Christina Obiero, Anna Turkova (Chair).

#### Section S2: Methods

##### Population pharmacokinetic modelling

Amoxicillin and clavulanate pharmacokinetic data were analysed using nonlinear mixed effects modelling in NONMEM v7.5.1 (ICON Development Solutions, Ellicott City, MD, USA) with the first-order conditional estimation with interaction (FOCE-I) method. Perl Speaks NONMEM v5.4.0, Pirana v24.9.0, and R v4.4.3 were used for data processing and visualisation.

Model development was conducted sequentially, with amoxicillin and clavulanate fitted independently before simultaneous modelling. For each compound, one- and two-compartment disposition models were investigated with first-order elimination. First-order absorption models were assessed with and without sequential lag time and zero-order input. Between-subject variability was tested on disposition parameters, and between-occasion variability on absorption parameters, with each dose defined as a separate occasion [1]. Residual unexplained variability was described using combined additive and proportional error models. Observations below the LOD were handled using the M7+ method [2].

Allometric scaling was included *a priori* on clearance and volume parameters with fixed exponents of 0.75 and 1, respectively, and total body weight or fat-free mass [3] were tested as body size descriptors. After accounting for allometry, we performed a stepwise covariate search with a forward inclusion of $P<0.05$ and backward elimination of $P<0.01$, considering also the stability of parameter estimates. A Hill function using postmenstrual age (PMA) was used to evaluate the maturation of amoxicillin and clavulanate clearance with age. To stabilise model estimates of the maturation function, parameter estimates were informed (using the PRIOR subroutine in NONMEM [4]) by Rhodin et al. [5], which describes the increase of glomerular filtration rate (GFR) from neonates to young adults. The prior estimates (relative standard errors [RSE%]) for the Hill coefficient and postmenstrual age at 50% maturation (PMA50) were 3.33 (6%) and 55.4 weeks (4%), respectively. Since individual values of gestational age at birth were unavailable, postmenstrual age was obtained using postnatal age and assuming a full gestation of 40 weeks. Glomerular filtration rate was estimated (eGFR) using the Schwartz formula [6] from participant serum creatinine concentrations and tested as a covariate on clearance. Weight-for-age z-score was investigated on absorption parameters and clearance. Additionally, amoxicillin/clavulanate formulation ratio, and amoxicillin or clavulanate dose were evaluated as covariates on amoxicillin and clavulanate absorption, bioavailability, and disposition parameters.

In the combined amoxicillin/clavulanate model, inclusion of correlations was investigated for between-subject and -occasion variability terms, and for the residual unexplained variability (using the L2 method in NONMEM). Model performance of intermediate and final models were assessed using goodness-of-fit diagnostics and visual predictive checks. Parameter precision of the final amoxicillin/clavulanate model was evaluated using a sampling importance resampling (SIR) procedure to derive 95% confidence intervals for all parameters [6].

#### Section S3: Pharmacokinetic model comparisons

Direct comparison of our results with previous studies is complicated by differences in the study cohort (body size, age/maturation, and clinical status), treatment (dose level and route of administration, i.e. oral vs intravenous [IV]), and model standardisation. To facilitate comparison, we used our model to predict CL/F and V/F for each previous study, applying the body size and age effects estimated in our model to the median participant characteristics reported for each study cohort (**Table S2 and S3**). Because our model is based on oral administration data only, our CL/F and V/F estimates cannot separate disposition (clearance, volume of distribution) from bioavailability (F). We therefore examined clearance and volume in parallel, reasoning that, for example, if our predictions are substantially higher than a comparison study for both CL/F and V/F, this pattern more likely reflects lower bioavailability (F) in our study than a true difference in disposition.

Predicted amoxicillin parameters were substantially higher than those from IV studies in both children (CL/F, 4.1-fold; V/F, 6.0-fold) [7], and adults (CL/F, 6.3-fold; V/F, 6.1-fold) [8], consistent with the expected effect of oral bioavailability on our apparent parameter estimates. Notably, our model also predicted consistently higher CL/F and V/F than the oral comparison studies, including in infants (CL/F, 3.4-fold; V/F, 2-fold) [9], where a difference in route of administration cannot explain the discrepancy. This may instead result from the higher amoxicillin dose level used in our study relative to all comparison studies. Given that amoxicillin bioavailability is reported to decrease at higher daily doses [10,11], this would lower F in our study and inflate our apparent CL/F and V/F. It implies that amoxicillin concentrations observed at the doses used here is lower than would be assumed under a linear, dose-proportional extrapolation. As a result, extrapolating from our amoxicillin model to the lower doses used in reference studies would underestimate concentrations in those studies. Volume was overpredicted more than clearance in the comparison with healthy adults (CL/F,

2- to 3-fold; V/F, 4-fold) [10,12], which is also plausible given that adults have a higher proportion of body fat than children [13]. Because amoxicillin and clavulanate are hydrophilic and distribute primarily into lean tissue rather than fat, scaling volume of distribution by total body weight is likely to overpredict adult volumes. When extrapolated to neonates, the discrepancy between our predictions and reported estimates was consistently greater for clearance than for volume of distribution (CL/F, 7-fold; V/F, 3-fold), suggesting that PMA-based maturation alone may not fully capture the physiological changes in clearance occurring around birth, as previously described [14].

Similar patterns were observed for clavulanate. Predicted CL/F and V/F exceeded IV comparisons in children (CL/F, 3.1-fold; V/F, 2.9-fold) [15] and adults (CL/F, 4.8-fold; V/F, 3.5-fold) [8], and among oral adult studies [12,16], volume was more overpredicted (2- to 3.2-fold) than clearance (1.4- to 1.8-fold), consistent with the amoxicillin comparisons above. In contrast, while amoxicillin CL/F and V/F were both overpredicted in neonates receiving suspension, clavulanate CL/F was similar to the comparison study (0.97-fold) and V/F was underpredicted (0.43-fold) [15]. Given that our model uses the same maturation function for both drugs, our clavulanate CL/F is likely still overestimated, as with amoxicillin. However, bioavailability in the comparison study may have been lower than in ours, owing to the known instability of clavulanate in suspension formulations [17]; a lower bioavailability (F) in that study would inflate its apparent CL/F and V/F, offsetting our CL/F overprediction and driving the apparent underprediction of V/F.

### Supplementary tables

#### Table S1. PediCAP trial dosing table for oral amoxicillin and amoxicillin/clavulanate (co-amoxiclav).

| Formulation | Weight band (kg) | # Tablets AM | # Tablets PM | Daily dose (mg) | Amoxicillin mg/kg range | Clavulanate mg/kg range |
|---|---|---|---|---|---|---|
| Amoxicillin (250mg tablets) | 3-<6 | 1 | 1 | 500 | 166.7–83.3 | |
| | 6-<10 | 2 | 1 | 750 | 125.0–75.0 | |
| | 10-<14 | 2 | 2 | 1000 | 100.0–71.4 | |
| | 14-<20 | 3 | 3 | 1500 | 107.1–75.0 | |
| | 20-<25 | 4 | 4 | 2000 | 100.0–80.0 | |
| | 25-<35 | 5 | 5 | 2500 | 100.0–71.4 | |
| Co-amoxiclav 7:1 (200/28.5 mg tablets) | 3-<6 | 1 | 1 | 400/57 | 133.3–66.7 | 19.0–9.5 |
| | 6-<10 | 2 | 2 | 800/114 | 133.3–80.0 | 19.0–11.4 |
| | 10-<14 | 2 | 3 | 1000/142.5 | 100.0–71.4 | 14.2–10.2 |
| | 14-<20 | 4 | 4 | 1600/228 | 114.3–80.0 | 16.3–11.4 |
| | 20-<25 | 5 | 5 | 2000/285 | 100.0–80.0 | 14.2–11.4 |
| | 25-<35 | 6 | 6 | 2400/342 | 96.0–68.6 | 13.7–9.8 |
| Co-amoxiclav 4:1 (250/62.5 mg tablets) | 3-<6 | 1 | 1 | 500/125 | 166.7–83.3 | 41.7–20.8 |
| | 6-<10 | 2 | 1 | 750/187.5 | 125.0–75.0 | 31.2–18.8 |
| | 10-<14 | 2 | 2 | 1000/250 | 100.0–71.4 | 25.0–17.9 |
| | 14-<20 | 3 | 3 | 1500/375 | 107.1–75.0 | 26.8–18.8 |
| | 20-<25 | 4 | 4 | 2000/500 | 100.0–80.0 | 25.0–20.0 |
| | 25-<35 | 5 | 5 | 2500/625 | 100.0–71.4 | 25.0–17.9 |
| Co-amoxiclav 14:1 (150/10.725 mg tablets) | 3-<6 | 2 | 1 | 450/32.175 | 150.0–75.0 | 10.7–5.4 |
| | 6-<10 | 3 | 2 | 750/53.625 | 125.0–75.0 | 8.9–5.4 |
| | 10-<14 | 4 | 3 | 1050/75.075 | 105.0–75.0 | 7.5–5.4 |
| | 14-<20 | 5 | 5 | 1500/107.25 | 107.1–75.0 | 7.7–5.4 |
| | 20-<25 | 7 | 6 | 1950/139.425 | 97.5–78.0 | 7.0–5.6 |
| | 25-<35 | 8 | 8 | 2400/171.6 | 96.0–68.6 | 6.9–4.9 |

#### Table S2. Pharmacokinetic comparisons of previously published studies on amoxicillin

| Parameter | Amoxicillin | | | | | | |
|---|---|---|---|---|---|---|---|
| Study | Current study | Keij FM, Schouwenburg S, et al. 2023 [18] | Barker CIS, et al. 2023 [9] | de Cock P, et al. 2015 [7] | Carlier M et al. 2013 [8] | de Velde F, et al. 2016 [10] | Witkowski G, et al. 1982 [12] |
| **Study Participants (n)** | Oral: 157 | Oral: 79 | Oral: 7 | IV: 50 | IV: 13 | Oral: 28 | Oral: 10 |
| | | IV: 182 | IV: 174 | | | | |
| **Study population** | Young children and infants, severe CAP | Preterm and term neonates | Young infants to adolescents, ICU / HDU | Young children and infants, ICU | Adults, ICU | Healthy adults | Healthy adults |
| **Route of administration** | Oral dispersible tablet | IV and oral suspension | IV and oral | IV | IV | Oral tablet | Oral tablet |
| **Median characteristics [a]** | | | | | | | |
| Weight (kg) | 8.4 | 3.6 | 3.8 | 14 | 75 | 77 | 67 |
| Postnatal age (years) | 1 | 0.0079 (2.9 days) | 0.14 (7.4 weeks) | 2.6 | 62 | 33 | 32 |
| Dose for study typical participant (mg/kg/day) | 91-98 | 75 | * | 1440-2016 | 53.3 | 9.7-23 | 7.5 |
| **Normalised CL/F (L/h)** | | | | | | | |
| Study model | 11.3 | 0.391 | 0.906 | 4.5 | 10.0 | 30.4 | 20.9 |
| Current model | 11.3 | 2.69 | 3.03 | 18.2 | 63.2 | 64.5 | 58.4 |
| **Normalised Vss/F (L/h)** | | | | | | | |
| Study model | 18.6 | 2.53 | 4.27 | 5.30 | 27.40 | 43.9 | 39.9 |
| Current model | 18.6 | 7.98 | 8.42 | 31.9 | 166 | 171 | 149 |

a. Based on the oral cohort of the study, if applicable
* Median oral dose was 24 mg/kg/dose. Dose per day and dosing interval not provided.
CL/F, apparent clearance. Vss/F, apparent volume of distribution at steady state

#### Table S3. Pharmacokinetic comparisons of previously published studies on clavulanate

| Parameter | Clavulanate | | | | | |
|---|---|---|---|---|---|---|
| Study | Current study | Schouwenburg S, Keij F, *et al.* 2024 [15] | de Cock P, *et al.* 2015 [7] | Carlier M, *et al.* 2013 [8] | de Velde F, *et al.* 2018 [16] | Witkowski G, *et al.* 1982 [12] |
| **Study Participants (n)** | Oral: 157 | Oral: 47<br>IV: 42 | IV: 50 | IV: 13 | Oral: 28 | Oral: 10 |
| **Study population** | Young children and infants, severe CAP | Neonates to infants, critically ill | Young children and infants, ICU | Adults, ICU | Healthy adults | Healthy adults |
| **Route of administration** | Oral dispersible tablet | IV and oral suspension | IV | IV | Oral tablet | Oral tablet |
| **Median characteristics** [a] | | | | | | |
| Weight (kg) | 8.4 | 3.6 | 14 | 75 | 77 | 67 |
| Postnatal age (years) | 1 | 0.0079 (2.9 days) | 2.6 | 62 | 33 | 32 |
| Dose for study typical participant (mg/kg/day) | 6.3-22 | 19 | 288-403 | 11 | 1.6 | 1.9 |
| **Normalised CL/F (L/h)** | | | | | | |
| Study model | 5.8 | 0.244 | 3.0 | 6.8 | 24.6 | 16.6 |
| Current model | 5.8 | 1.41 | 9.4 | 32.7 | 33.3 | 30.2 |
| **Normalised Vss/F (L/h)** | | | | | | |
| Study model | 7.35 | 7.11 | 4.41 | 19.20 | 33 | 18.4 |
| Current model | 7.35 | 3.15 | 12.6 | 65.6 | 67.4 | 59.0 |

a. Based on the oral cohort of the study, if applicable.
CL/F, apparent clearance. Vss/F, apparent volume of distribution at steady state

### Supplementary figures

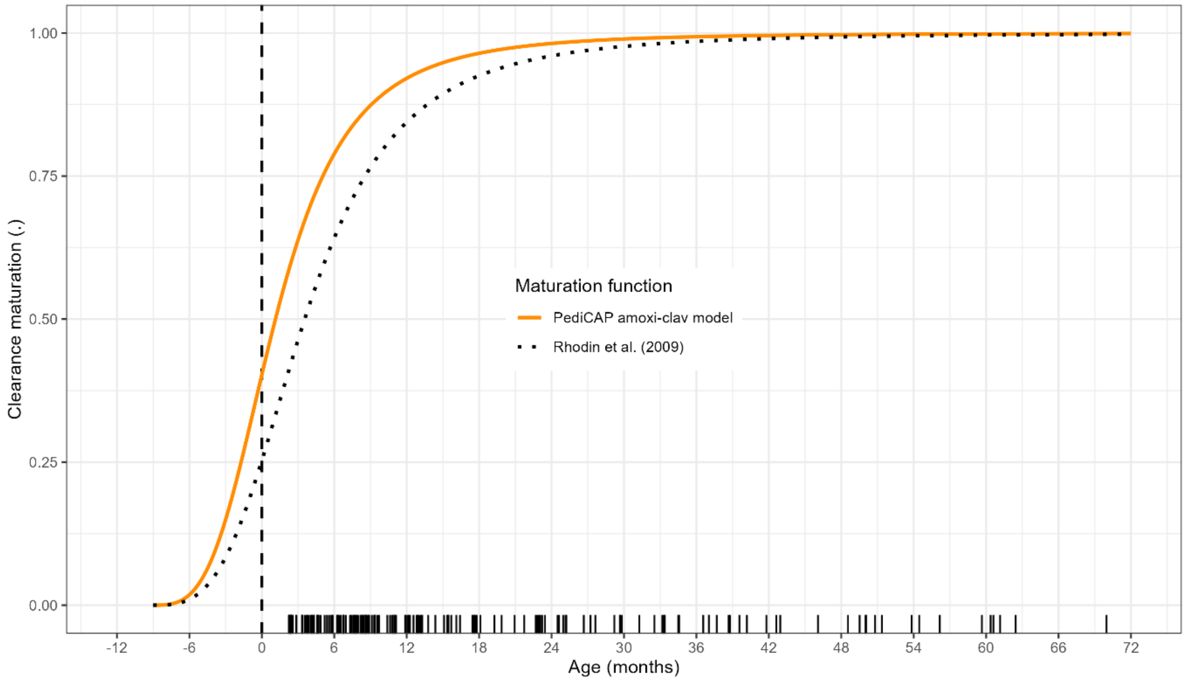

**Figure S1. Clearance maturation profile.** Clearance maturation is plotted against postnatal age (months) using our model maturation function for amoxicillin/clavulanate (solid orange line; Hill = 3.42, PMA50 = 44.9 weeks) and the glomerular filtration rate maturation function reported by Rhodin et al. [5] (dotted black line; Hill = 3.33, PMA50 = 55.4 weeks). Birth at 40 weeks gestational age is indicated by the vertical dashed line. Black rug marks along the x-axis indicate the observed postnatal ages in our dataset.
